# Combining Real and Synthetic Data to Overcome Limited Training Datasets in Multimodal Learning

**DOI:** 10.1101/2025.07.16.25331662

**Authors:** Niccolo Marini, Zhaohui Liang, Sivaramakrishnan Rajaraman, Zhiyun Xue, Sameer Antani

**Affiliations:** Division of Intramural Research, National Library of Medicine, National Institutes of Health Bethesda, MD, 290894, USA

## Abstract

Biomedical data are inherently multimodal, capturing complementary aspects of a patient condition. Deep learning (DL) algorithms that integrate multiple biomedical modalities can significantly improve clinical decisionmaking, especially in domains where collecting data is not simple and data are highly heterogeneous. However, developing effective and reliable multimodal DL methods remains challenging, requiring large training datasets with paired samples from modalities of interest. An increasing number of de-identifed biomedical datasets are publicly accessible, though they still tend to be unimodal. For example, several publicly available skin lesion datasets aid automated dermatology clinical decision-making. Still, they lack annotated reports paired with the images, thereby limiting the advance and use of multimodal DL algorithms. This work presents a strategy exploiting real and synthesized data in a multimodal architecture that encodes finegrained text representations within image embeddings to create a robust representation of skin lesion data. Large language models (LLMs) are used to synthesize textual descriptions from image metadata that are subsequently paired with the original skin lesion images and used for model development. The architecture is evaluated on the classification of skin lesion images, considering nine internal and external data sources. The proposed multimodal representation outperforms the unimodal one on the classification of skin lesion images, achieving superior performance in every tested dataset.

## 1. Introduction

The increasing availability of multimodal biomedical data [25] is driving the development of novel deep learning (DL) algorithms aimed at analyzing and representing complex medical information, particularly in domains characterized by limited annotated data and high intrinsic heterogeneity in terms of diseases, such as skin lesion classification from dermatology images.

In routine clinical practice, the use of multiple biomedical modalities aids in the capture of complementary aspects of the patient’s health and facilitates the early identification of potentially dangerous diseases [1, 23]. For example, biomedical images provide low-level visual features about disease manifestations, while associated textual reports summarize high-level diagnostic findings identified during the sample examination [14]. Multimodal learning focuses on training DL models integrating samples from various modalities to capture the relationships across different sources of information and to improve the data representations. Therefore, the integration of multiple information sources is particularly advantageous in fields where collecting annotated datasets is time-consuming and data are inherently heterogeneous [24]. Furthermore, the adoption of multimodal learning algorithms is also rapidly increasing [22], helped by the increasing amount of data collected by healthcare systems [1].

However, a significant challenge in the development of multimodal algorithms is that they require sufficiently large datasets with paired samples of the modalities under consideration. Training datasets including paired samples corresponding to the same clinical case are necessary to allow models to associate features across modalities and then to learn cross-modal relationships [2]. An additional complexity is the use of large architectures that are needed to capture relationships among modalities [1, 11]. Large datasets are also necessary to guarantee the robustness and the generalization of multimodal algorithms [16] on unseen data, due to the high dimensionality and complexity of biomedical data, especially considering unstructured samples like images and textual reports. Multimodal learning models, such as foundation models [11], usually include large architectures to capture fine-grained relationships among modalities and therefore require to be trained with large training datasets [16], to avoid overfitting.

Even if the collection of multimodal biomedical data is increasing, some biomedical domains, such as dermatology, still lack large datasets that include samples from multiple modalities. Dermatology focuses on the acquisition and analysis of dermatological skin lesions to identify morphological patterns and features indicative of disease [27], that can be malignant and dangerous (skin cancer is the most common cancer in the United States of America [9, 10]). Despite the large amount of publicly available resources including dermatology images, publicly available datasets lack a large collection of paired samples including images and texts, preventing the development of multimodal architectures. To the best of our knowledge, only a single publication presented a multimodal model that combined dermatology images and reports [29]. However, it includes a private dataset, limiting advances in the field. The metadata linked to images in the publicly available datasets can be exploited to build reports through the use of Large Language Models (LLM) [13]. The large diffusion of LLMs allows both image analysis and automatic generation of reports, but the vast majority of LLMs are not built to analyze medical data and may produce hallucinations, depending on the specific prompt adopted to generate the reports.

The goal of this paper is to exploit image metadata and LLMs to synthesize the corresponding report to train a multimodal architecture, analyzing images and reports. Furthermore, considering the limited amount of training data, annotations are exploited to enrich the data representation. Our proposed architecture generates a robust shared dermatology representation, encoding fine-grained textual concepts within image representations. The transfer of information from reports (highly informative) to images (that are less informative) is known as multimodal co-learning [15], which helps to build more accurate robust data representations [19]. The architecture is tested on the classification of skin lesion images, considering five classes, comparing the performance of the architecture trained only with images and the architecture trained combining information from both modalities.

## 2. Methods

### 2.1. Multimodal architecture

Our proposed multimodal architecture includes two branches to analyze both modalities and a specific training strategy to integrate visual and textual information from dermatology data (paired images and textual reports). It consists of two modality-specific encoders with a shared backbone, including a joint classifier. Here, the encoders produce fixed-size embeddings which feed the shared classifier that processes both types of embeddings. The image encoder is a CNN (i.e. DenseNet121, pre-trained using sim-CLR algorithm [4]) and the text encoder is a PubMed Bert [8] (pre-trained on PubMed®data). The joint classifier is trained to optimize the loss function (i.e. Cross-Entropy) on the class predictions for both images and reports. The choice to share the classifier weights for both modalities is part of the strategy designed to align the modalities. It aids in combining loss functions and the shared classifier weights. Three loss functions are applied to the feature embeddings from both modalities: L1-loss function, Cosine Similarity and a self-supervised (SSL) algorithm (NT-Xent and InfoNCE [17] are tested). Considering the limited amount of training samples, the use of shared weights aims to smoothly align the modalities and to avoid a possible overfitting caused by the three loss functions. The architecture is trained considering two setups, unimodal (i.e. trained with only images), multimodal (i.e. trained with images and reports), while it is evaluated only on the image classification errors (Cross-Entropy loss) during the validation. Figure 1 provides an overview of the multimodal architecture.

**Figure 1.**
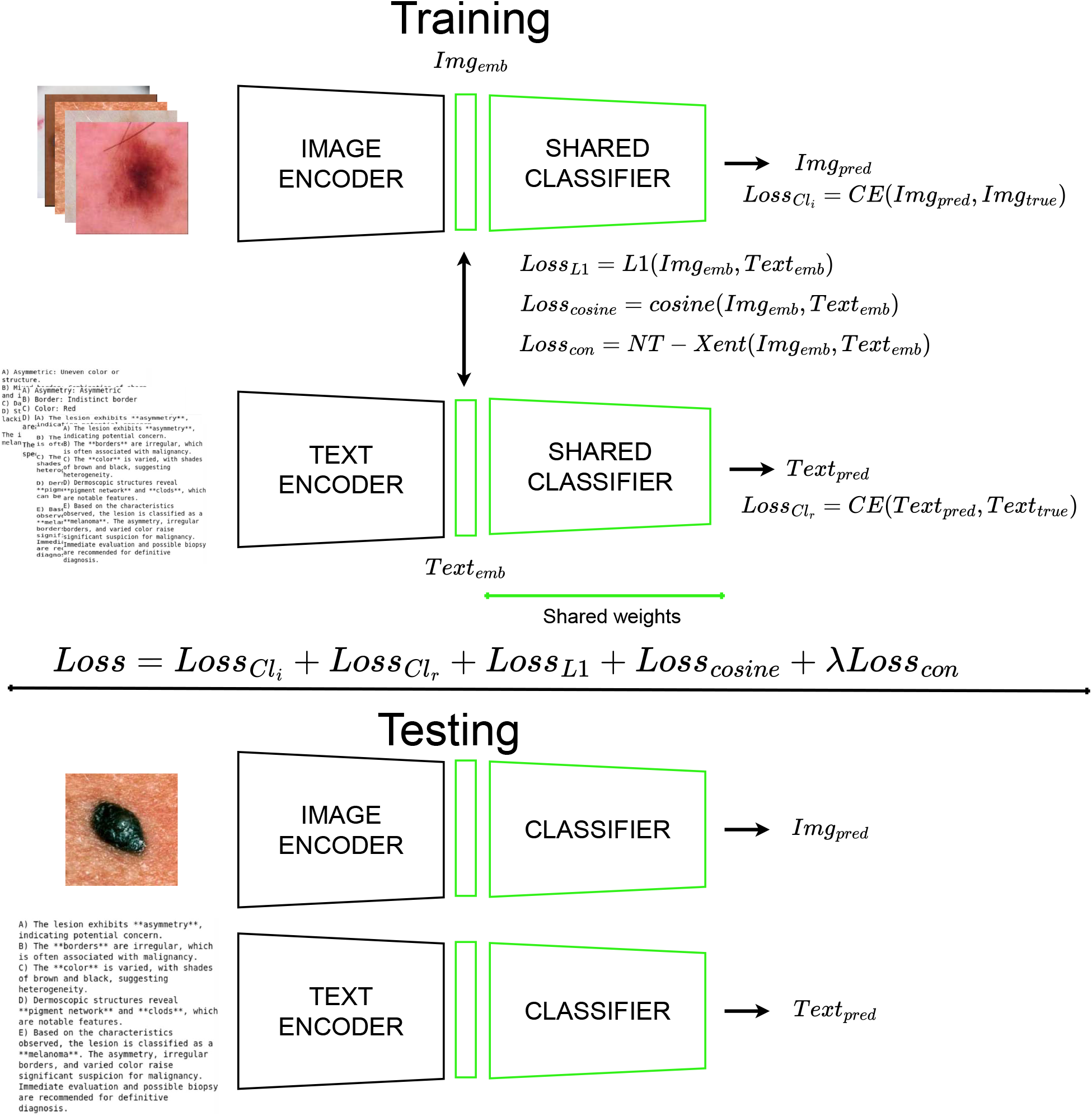
Overview of the multimodal architecture, including with two input branches for skin lesion images and textual reports, followed by a shared projection head and classifier. The training phase requires both modalities with a composite loss combining classification, NT-Xent, L1, and cosine similarity terms to align image and text representations. At inference, each modality can be used independently.

### 2.2. Datasets

The data used to develop the architecture include images and textual reports, collected from multiple sources and split into three partitions (i.e. training, validation and testing).

The architecture is trained using both images and reports, while it is tested using only images. The test partition includes only images because the goal of the architecture is to transfer information from reports, that are highly informative, to images that are less informative. Furthermore, only the performance on images is clinically-relevant: a possible performance increase on the assessment of textual reports does not show any advantage in clinical practice, where images are analyzed by experts, that subsequently produce a report.

Data are collected from multiple medical sources to guarantee heterogeneity, in terms of the visual appearance of a disease manifestation, and to test the robustness of the architecture to generalize on data collected from medical sources that are different from the ones used to train the model. Dermatology images include both dermoscopic images [3] (i.e., high-resolution, magnified photographs of skin lesions captured using polarized or non-polarized light) and clinical pictures (i.e., images including skin lesions, usually a non-magnified and non-centered image). Reports are generated from the image metadata, as shown in 2.3.

Data are split in three partitions, as shown in Table 1.

**Table 1.** Composition of the dataset collected from multiple sources to guarantee lesion heterogeneity. The dataset includes skin lesions and the synthesized reports and it is split into training, validation and testing partitions. The dataset includes five classes: Benign Keratosis (BEK), Benign Nevus (NEV), Actinic Keratosis (ACK), Basal-Cell Cancer (BCC), Melanoma (MEL).

| Dataset | BEK | NEV | ACK | BCC | MEL | Tot |
| --- | --- | --- | --- | --- | --- | --- |
| **Training partition** |  |  |  |  |  |  |
| BCN20000 | 796 | 2944 | 515 | 1966 | 1999 | 8220 |
| Derm12345 | 430 | 1683 | 16 | 291 | 263 | 2683 |
| DDI | 22 | 22 | 0 | 7 | 5 | 56 |
| Derm7pt | 46 | 18 | 0 | 26 | 160 | 250 |
| DermNet | 171 | 39 | 38 | 133 | 62 | 443 |
| Total | 1465 | 4706 | 569 | 2423 | 2489 | 11652 |
| **Validation partition** |  |  |  |  |  |  |
| BCN20000 | 170 | 630 | 110 | 421 | 428 | 1759 |
| Derm12345 | 110 | 455 | 13 | 47 | 55 | 680 |
| Derm7pt | 21 | 8 | 0 | 12 | 60 | 110 |
| DermNet | 25 | 7 | 6 | 19 | 10 | 67 |
| Total | 326 | 1100 | 129 | 499 | 562 | 2616 |
| **Testing partition** |  |  |  |  |  |  |
| BCN20000 | 172 | 632 | 112 | 422 | 430 | 1768 |
| Derm12345 | 135 | 534 | 8 | 85 | 82 | 844 |
| Derm7pt | 13 | 5 | 0 | 7 | 45 | 70 |
| DermNet | 49 | 11 | 11 | 38 | 18 | 127 |
| HAM10000 | 1338 | 7737 | 378 | 622 | 1305 | 11380 |
| SKINL2 | 33 | 97 | 0 | 40 | 28 | 198 |
| Fitzpatrick17k | 30 | 39 | 5 | 36 | 115 | 225 |
| Buenos Aires | 88 | 602 | 63 | 340 | 253 | 1346 |
| SD198 | 60 | 32 | 62 | 13 | 38 | 205 |
| PAD UFES 20 | 52 | 68 | 13 | 64 | 26 | 223 |
| Total | 1970 | 9757 | 651 | 1667 | 2340 | 16386 |

The splitting aims to guarantee that training and validation partitions include both dermoscopy and clinical images. Furthermore, the testing partition includes external datasets (i.e. different source than the ones used for training) to assess how the model generalizes on unseen data. The training and validation partitions include data from BCN20000 (dermoscopy, D), Derm12345 [28] (D), Derm7pt [12] (C), DermNet ^1^ (clinical, C), DDI [6] (C); the testing partitions include data from BCN20000, Derm7pt, DermNet, derm12345, DDI, HAM10000 [26] (D), SKINL2 [7] (D), Fitzpatrick17k^2^ (C), Hospital Italiano Buenos Aires [20] (D), SD198^3^ (C), PAD UFES 20 [18] (C). The original datasets include many images, but only a subset of the images are selected, corresponding to the following classes: Benign Keratosis (BEK, benign), Benign Nevus (NEV, benign), Actinic Keratosis (ACK, pre-malignant), Basal-Cell Cancer (BCC, malignant), Melanoma (MEL, malignant).

### 2.3. Report generation

The reports are generated by exploiting metadata related to the selected input images. Metadata include information about the class of lesion included in the image, such as melanocytic nevus or melanoma. Some metadata include information that can be linked to one of the five classes selected within the problem (e.g., Melanocytic Nevus is a subclass for Benign nevus, Bowen’s disease is a subclass for Actinic Keratosis). Considering the heterogeneous structure of the metadata, two types of input reports are generated: automatically structured generated and synthesized reports. The automatically structured reports are generated filling a string of text: “The image includes a *bening* / *malignant* skin lesion, specifically a *class* (specifically a *subclass*), where *bening* / *malignant, class, subclass* are information collected from the metadata (*subclass* only if available, otherwise it is not added to the report). The synthesized reports are generated using an LLM (i.e. gpt-4o-mini was used). The starting point for the generation is the automatically structured reports, that are filled with additional fields related to the structure of the skin, the color of the image, some dermoscopic structures, and the symmetry/asymmetry of the lesion. Each one of the additional fields include a set of options to be selected, to avoid hallucinations from the LLMs, that are not trained on the specific knowledge requested to analyze dermatology data.

### 2.4. Experimental setup

The experimental setup involves hyperparameter optimization, data encoding, the loss functions adopted to align modalities and data augmentation strategies, and the strategy to handle the class unbalance in the training partition. The hyperparameters have been chosen with a grid search algorithms, focusing on number of epochs (10), learning rate (1e-4), optimizer (Adam), batch size (32), decay weight (1e-5), loss functions weight (1 for all the losses, except the SSL loss functions, which is 0.5 when NT-Xent is used, 0.25 when InfoNCE is used), the temperature of the SSL loss functions (0.5 for NT-Xent, 0.07 for InfoNCE). The data encoding involved the branches to process data modalities: the main experimental choice involves the image encoder, selecting a CNN architecture (i.e. DenseNet121). The choice of a CNN is driven by multiple aspects linked to the need to avoid overfitting: the lack of pre-trained Visual Transformers (ViT) and the limited number of samples cannot guarantee to learn a good image representation on the pre-trained image backbone, in contrast to pre-trained CNN (refined with simCLR pre-training) that can achieve that. The loss functions adopted to align modalities include three elements: L1-loss, Cosine Loss and an SSL (Nt-Xent or InfoNCE) loss function. The first two loss functions are adopted to match the feature embeddings. However, the functions match the representations samplewise within a batch. Therefore, another loss function is adopted (i.e. the SSL loss function). Among the possible choices, NT-Xent and InfoNCE loss were compared. The data augmentation strategy involves both the augmentation of images and reports. The image augmentation strategy includes rotations, horizontal and vertical flipping and a light RGB color augmentation. The report augmentation includes the turnover between the two types of reports, presented in 2.3. Classwise data augmentation is applied to balance the training dataset: samples from less represented classes are selected more frequently, providing their augmented versions to the model.

## 3. Results

The multimodal architecture outperforms the unimodal baselines in the classification of dermatology images for both internal and external testing partitions and leads to a more robust representation.

Table 2 summarizes the results, reporting the average and standard deviation of Cohen’s *κ*-score across ten independent model repetitions, for every training setup (i.e. unimodal, multimodal, MM). Considering that the data were collected from medical sources different than the ones used to train the model, the results the robustness of the multimodal learning strategy to data distribution shifts.

**Table 2.** Performance of the proposed multimodal architecture on skin lesion classification across internal and external test sets. Results include unimodal and multimodal models (with NT-Xent and InfoNCE losses), evaluated via Cohen’s *κ*-score. Statistically significant differences (Wilcoxon test) are marked with (*).

| Dataset | Unimodal | MM (NT-Xent) | MM (InfoNCE) |
| --- | --- | --- | --- |
| **Internal partition** |  |  |  |
| **BCN20000** | $0.675 \pm 0.015$ | **$0.705 \pm 0.021^*$** | $0.697 \pm 0.025^*$ |
| **Derm12345** | $0.731 \pm 0.025$ | $0.724 \pm 0.013$ | **$0.743 \pm 0.013$** |
| **Derm7pt** | $0.591 \pm 0.052$ | **$0.609 \pm 0.041$** | $0.604 \pm 0.066$ |
| **DermNet** | $0.706 \pm 0.049$ | $0.723 \pm 0.044$ | **$0.728 \pm 0.041$** |
| **External partition** |  |  |  |
| **HAM10000** | $0.470 \pm 0.021$ | $0.483 \pm 0.017$ | **$0.490 \pm 0.012^*$** |
| **SKINL2** | $0.722 \pm 0.052$ | **$0.739 \pm 0.068$** | $0.720 \pm 0.058$ |
| **Fitzpatrick17k** | $0.446 \pm 0.031$ | $0.463 \pm 0.036$ | **$0.464 \pm 0.046$** |
| **Buenos Aires** | $0.415 \pm 0.065$ | $0.422 \pm 0.038$ | **$0.442 \pm 0.038$** |
| **SD198** | $0.492 \pm 0.048$ | **$0.550 \pm 0.047$** | $0.499 \pm 0.055$ |
| **PAD UFES 20** | $0.361 \pm 0.056$ | **$0.385 \pm 0.050$** | $0.374 \pm 0.054$ |

Both SSL loss functions adopted in the multimodal training strategy achieve higher performance than the unimodal network, although neither emerges as the more robust of the two.

Table 3 summarizes the evaluation of the Silhouette score [21], a metric to assess the quality of a feature representation by measuring how well each sample is clustered, reflecting both intra-cluster cohesion and inter-cluster separation concerning the classes. From the results it is evident that the application of the multimodal training strategy leads to higher scores. Consequently, the multimodal architecture better separates sample classes.

**Table 3.**
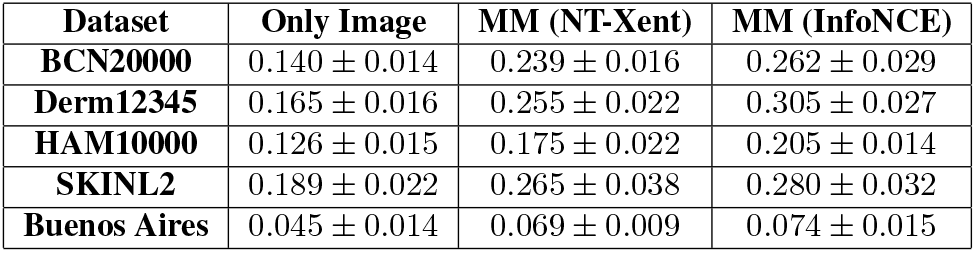
Overview on the silhouette score evaluated on the test partitions including dermascopies. The evaluation involves the unimodal and the multimodal representation (considering both NTXent and InfoNCE as SSL loss function), reporting the average and the standard deviation of the ten models trained.

| Dataset | Only Image | MM (NT-Xent) | MM (InfoNCE) |
| --- | --- | --- | --- |
| **BCN20000** | $0.140 \pm 0.014$ | $0.239 \pm 0.016$ | $0.262 \pm 0.029$ |
| **Derm12345** | $0.165 \pm 0.016$ | $0.255 \pm 0.022$ | $0.305 \pm 0.027$ |
| **HAM10000** | $0.126 \pm 0.015$ | $0.175 \pm 0.022$ | $0.205 \pm 0.014$ |
| **SKINL2** | $0.189 \pm 0.022$ | $0.265 \pm 0.038$ | $0.280 \pm 0.032$ |
| **Buenos Aires** | $0.045 \pm 0.014$ | $0.069 \pm 0.009$ | $0.074 \pm 0.015$ |

## 4. Discussion & Conclusion

Since biomedical data are inherently multimodal, DL algorithms that integrate multiple modalities can significantly improve clinical decision-making. Even though a growing number of biomedical data sets are becoming available for use with AI algorithms, they tend to be unimodal. We demonstrate the effectiveness of multimodal learning through use of several publicly available skin lesion datasets. However, since they lack annotated reports paired with the images, we present a strategy exploiting real and LLM synthesized data in a multimodal architecture that encodes fine-grained text representations within image embeddings to create a robust representation of skin lesion data.

The results demonstrate that our multimodal training strategy significantly improves classification performance compared to unimodal approaches. This improvement can be attributed to the ability of the model to transfer meaningful information embedded within the textual reports to the image representations. The model benefits from meaningful report information, even though it is synthesized, highlighting the advantages of co-learning strategies in the domain of dermatology, where lesions are highly heterogeneous and annotated data may be scarce. The performance gains were consistent across both internal and external testing partitions, and also considering different imaging types, including dermoscopic and clinical photographs, which are inherently noisier. The findings are also confirmed through the use of the silhouette score, which shows that multimodal representations are more compact and the class dispersion is lower.

Despite the overall performance improvement, the results show considerable variability, as indicated by relatively high standard deviations across experiments, that is not alleviated by the multimodal learning strategy. One possible explanation can be the overfitting on training data, which is only partially addressed by the architectural design.

The proposed multimodal strategy shows great potential in scenarios where large, well-curated datasets with paired modalities are not available. It offers a possibility to bridge the knowledge gap between modalities, allowing models to leverage cross-modal information even under suboptimal data conditions. Future work will focus on further stabilizing the training process and exploring strategies to better align representations across modalities to reduce performance variability and how to exploit the common representation to link knowledge across modalities, without minimal need for strictly annotated data.

## Data Availability

All datasets are publicly available.
BCN20000: https://figshare.com/articles/journal_contribution/BCN20000_Dermoscopic_Lesions_in_the_Wild/24140028/1
dermnet12345: https://api.isic-archive.com/doi/derm12345/
DermNet: https://www.kaggle.com/datasets/shubhamgoel27/dermnet
Derm7pt: https://derm.cs.sfu.ca/Download.html
HAM10000: https://dataverse.harvard.edu/dataset.xhtml?persistentId=doi:10.7910/DVN/DBW86T
SKINL2: https://www.it.pt/AutomaticPage?id=3459
Fitzpatrick17k: https://github.com/mattgroh/fitzpatrick17k
Hospital_Italiano_Buenos_Aires: https://api.isic-archive.com/collections/251/
PAD_UFES_20: https://data.mendeley.com/datasets/zr7vgbcyr2/1
SD198: https://huggingface.co/datasets/resyhgerwshshgdfghsdfgh/SD-198

## Acknowledgments

This work is supported by the Intramural Research Program of the National Library of Medicine (NLM), part of the U.S. National Institutes of Health (NIH).

## Footnotes

1 https://www.kaggle.com/datasets/shubhamgoel27/dermnet. Retrieved April 28, 2025

2 https://github.com/mattgroh/fitzpatrick17k, Retrieved April 28, 2025

3 https://huggingface.co/datasets/resyhgerwshshgdfghsdfgh/SD-198, Retrieved April 28, 2025

## References

[1] Julián N Acosta, Guido J Falcone, Pranav Rajpurkar, and Eric J Topol. Multimodal biomedical ai. Nature medicine, 28(9):1773–1784, 2022. 2

[2] Rawan AlSaad, Alaa Abd-Alrazaq, Sabri Boughorbel, Arfan Ahmed, Max-Antoine Renault, Rafat Damseh, and Javaid Sheikh. Multimodal large language models in health care: applications, challenges, and future outlook. Journal of medical Internet research, 26:e59505, 2024. 2

[3] M Emre Celebi, Noel Codella, and Allan Halpern. Dermoscopy image analysis: overview and future directions. IEEE journal of biomedical and health informatics, 23(2): 474–478, 2019. 4

[4] Ting Chen, Simon Kornblith, Mohammad Norouzi, and Geoffrey Hinton. A simple framework for contrastive learning of visual representations. In International conference on machine learning, pages 1597–1607. PmLR, 2020. 2, 3

[5] Marc Combalia, Noel CF Codella, Veronica Rotemberg, Brian Helba, Veronica Vilaplana, Ofer Reiter, Cristina Carrera, Alicia Barreiro, Allan C Halpern, Susana Puig, et al. Bcn20000: Dermoscopic lesions in the wild. arXiv preprint 1908.02288, 2019. 4

[6] Roxana Daneshjou, Kailas Vodrahalli, Weixin Liang, Roberto A Novoa, Melissa Jenkins, Veronica Rotemberg, Justin Ko, Susan M Swetter, Elizabeth E Bailey, Olivier Gevaert, et al. Disparities in dermatology ai: Assessments using diverse clinical images. arXiv preprint 2111.08006, 2021. 4

[7] Sergio MM de Faria, Jose N Filipe, Pedro MM Pereira, Luis MN Tavora, Pedro AA Assuncao, Miguel O Santos, Rui Fonseca-Pinto, Felicidade Santiago, Victoria Dominguez, and Martinha Henrique. Light field image dataset of skin lesions. In 2019 41st Annual International Conference of the IEEE Engineering in Medicine and Biology Society (EMBC), pages 3905–3908. IEEE, 2019. 4

[8] Yu Gu, Robert Tinn, Hao Cheng, Michael Lucas, Naoto Usuyama, Xiaodong Liu, Tristan Naumann, Jianfeng Gao, and Hoifung Poon. Domain-specific language model pretraining for biomedical natural language processing. ACM Transactions on Computing for Healthcare (HEALTH), 3(1): 1–23, 2021. 2

[9] Gery P Guy Jr, Steven R Machlin, Donatus U Ekwueme, and K Robin Yabroff. Prevalence and costs of skin cancer treatment in the us, 2002-2006 and 2007-2011. American journal of preventive medicine, 48(2):183–187, 2015. 2

[10] Gery P Guy Jr, Cheryll C Thomas, Trevor Thompson, Meg Watson, Greta M Massetti, Lisa C Richardson, Centers for Disease Control, Prevention (CDC), et al. Vital signs: melanoma incidence and mortality trends and projections-united states, 1982-2030. MMWR Morb Mortal Wkly Rep, 64(21):591–596, 2015. 2

[11] Yuting He, Fuxiang Huang, Xinrui Jiang, Yuxiang Nie, Minghao Wang, Jiguang Wang, and Hao Chen. Foundation model for advancing healthcare: challenges, opportunities and future directions. IEEE Reviews in Biomedical Engineering, 2024. 2

[12] Jeremy Kawahara, Sara Daneshvar, Giuseppe Argenziano, and Ghassan Hamarneh. Seven-point checklist and skin lesion classification using multitask multimodal neural nets. IEEE journal of biomedical and health informatics, 23(2): 538–546, 2018. 4

[13] Zhuoyan Li, Hangxiao Zhu, Zhuoran Lu, and Ming Yin. Synthetic data generation with large language models for text classification: Potential and limitations. arXiv preprint 2310.07849, 2023. 2

[14] Weimin Lyu, Xinyu Dong, Rachel Wong, Songzhu Zheng, Kayley Abell-Hart, Fusheng Wang, and Chao Chen. A multimodal transformer: Fusing clinical notes with structured ehr data for interpretable in-hospital mortality prediction. In AMIA Annual Symposium Proceedings, page 719, 2023. 2

[15] Niccoló Marini Stefano Marchesin, Marek Wodzinski, Alessandro Caputo, Damian Podareanu, Bryan Cardenas Guevara, Svetla Boytcheva, Simona Vatrano, Filippo Fraggetta, Francesco Ciompi, et al. Multimodal representations of biomedical knowledge from limited training whole slide images and reports using deep learning. Medical Image Analysis, 97:103303, 2024. 2

[16] Michael Moor, Oishi Banerjee, Zahra Shakeri Hossein Abad, Harlan M Krumholz, Jure Leskovec, Eric J Topol, and Pranav Rajpurkar. Foundation models for generalist medical artificial intelligence. Nature, 616(7956):259–265, 2023. 2

[17] Aaron van den Oord, Yazhe Li, and Oriol Vinyals. Representation learning with contrastive predictive coding. arXiv preprint 1807.03748, 2018. 3

[18] Andre GC Pacheco, Gustavo R Lima, Amanda S Salomao, Breno Krohling, Igor P Biral, Gabriel G de Angelo, Fábio CR Alves Jr, José GM Esgario, Alana C Simora, Pedro BC Castro, et al. Pad-ufes-20: A skin lesion dataset composed of patient data and clinical images collected from smartphones. Data in brief, 32:106221, 2020. 4

[19] Anil Rahate, Rahee Walambe, Sheela Ramanna, and Ketan Kotecha. Multimodal co-learning: Challenges, applications with datasets, recent advances and future directions. Information Fusion, 81:203–239, 2022. 2

[20] María Agustina Ricci Lara, María Victoria Rodríguez Kowalczuk, Maite Lisa Eliceche, María Guillermina Ferraresso, Daniel Roberto Luna, Sonia Elizabeth Benitez, and Luis Daniel Mazzuoccolo. A dataset of skin lesion images collected in argentina for the evaluation of ai tools in this population. Scientific Data, 10(1):712, 2023. 4

[21] Peter J Rousseeuw. Silhouettes: a graphical aid to the interpretation and validation of cluster analysis. Journal of computational and applied mathematics, 20:53–65, 1987. 5

[22] Thanveer Shaik, Xiaohui Tao, Lin Li, Haoran Xie, and Juan D Velásquez. A survey of multimodal information fusion for smart healthcare: Mapping the journey from data to wisdom. Information Fusion, 102:102040, 2024. 2

[23] Sören Richard Stahlschmidt, Benjamin Ulfenborg, and Jane Synnergren. Multimodal deep learning for biomedical data fusion: a review. Briefings in bioinformatics, 23(2):bbab569, 2022. 2

[24] Zhaoyi Sun, Mingquan Lin, Qingqing Zhu, Qianqian Xie, Fei Wang, Zhiyong Lu, and Yifan Peng. A scoping review on multimodal deep learning in biomedical images and texts. Journal of biomedical informatics, 146:104482, 2023. 2

[25] Amalio Telenti and Xiaoqian Jiang. Treating medical data as a durable asset. Nature Genetics, 52(10):1005–1010, 2020. 2

[26] Philipp Tschandl, Cliff Rosendahl, and Harald Kittler. The ham10000 dataset, a large collection of multi-source dermatoscopic images of common pigmented skin lesions. Scientific data, 5(1):1–9, 2018. 4

[27] Richard B Weller, Hamish JA Hunter, and Margaret W Mann. Clinical dermatology. John Wiley & Sons, 2014. 2

[28] Abdurrahim Yilmaz, Sirin Pekcan Yasar, Gulsum Gencoglan, and Burak Temelkuran. Derm12345: A large, multisource dermatoscopic skin lesion dataset with 40 subclasses. Scientific Data, 11(1):1302, 2024. 4

[29] Juexiao Zhou, Xiaonan He, Liyuan Sun, Jiannan Xu, Xiuying Chen, Yuetan Chu, Longxi Zhou, Xingyu Liao, Bin Zhang, Shawn Afvari, et al. Pre-trained multimodal large language model enhances dermatological diagnosis using skingpt-4. Nature Communications, 15(1):5649, 2024. 2

